# Machine Learning for Paediatric Related Decision Support in Emergency Care – A UK and Ireland Network Survey Study of Emergency Staff

**DOI:** 10.1101/2025.07.06.25330977

**Authors:** Fiona Leonard, Mark D Lyttle, Dympna O’Sullivan, John Gilligan, Damian Roland, Michael Barrett, PERUKI

**Author notes:** Corresponding author: (FL).

## Abstract

There is great potential for artificial Intelligence (AI) and machine learning (ML) to support decision making in emergency departments (ED), however their implementation in routine clinical practice remains limited. The objective of this study was to assess the understanding, experience and perspectives of the wider paediatric ED workforce (nurses, doctors and support staff) in the United Kingdom and Ireland on the use of ML decision support tools. A voluntary and anonymised survey was carried out across 75 sites. The survey consisted of questions on self-reported knowledge of AI concepts (which included watching a short video), exposure, barriers to adoption, training and potential ML applications. Mostly quantitative analysis was performed. The survey had a 72.3% response rate (660 responses). Prior to viewing the video, understanding of AI concepts varied, with AI the most understood (60.3%) compared to deep learning at 19.1%. Post-video, 40.2% of respondents changed their answers. While many respondents had experience of decision rule-based systems (53.3%), only 7.7% reported using ML based tools. Key barriers to adoption included uncertainty about suitable clinical applications (42.8%), lack of skilled resources (example: data scientists or engineers) (37.0%), limited model explainability (31.5%) and poor data quality (28.8%). Many respondents believed these tools could integrate well into clinical workflows (60.7%), would trust these tools (58.2%), and had a strong interest in furthering their knowledge in ML (78.2%). Early warning systems and radiology applications ranked highest, and diagnosis of mental health conditions lowest, for doctors and nurses. These findings show knowledge gaps and limited exposure to ML tools, yet a strong interest in learning is evident. To realise the potential of ML in children’s emergency care, domain specific AI literacy, improved model transparency, investment in infrastructure and resources, and better integration into clinical workflows are essential.

**Author Summary:** In this study we set out to understand how the staff (nurses, doctors, and support staff) in children’s emergency care across the United Kingdom and Ireland perceive and engage with machine learning decision support tools. Whilst much research into these tools exists, few are used in clinical practice. To understand why, we conducted a multi-centre survey, asking participants about their understanding of artificial intelligence concepts, past use of machine learning tools, barriers to adoption, and views on the most useful clinical applications. We found that although familiarity with machine learning and its application was low, interest was high, with many believing these tools could be integrated well into clinical workflows. Early warning systems and radiology were viewed as the most promising uses. Barriers included uncertainty around where machine learning could add value, not understanding how these tools generated their output, lack of skilled resources to implement these tools, and concerns around poor data quality. Our findings on the lack of artificial intelligence literacy align with surveys internationally. Addressing these challenges, along with providing targeted training tailored to each role, will be key to supporting safe, confident, and effective use of machine learning tools for decision support in children’s emergency care.

## Introduction

There has been a rapid increase in research exploring the use of artificial intelligence (AI) and machine learning (ML) in emergency departments (ED), with many studies demonstrating useful decision support applications. However despite promising results, relatively few have progressed beyond the research stage to be implemented into routine clinical practice [1]. One contributing factor is a lack of trust, not only in the AI tools themselves but across the full lifecycle, including their design, validation and implementation [2]. Understanding perspectives of frontline staff is critical in identifying barriers to adoption, as well as workforce readiness to embrace AI-driven technologies for decision support.

Clinical decision support system tools can be grouped into knowledge-based and non-knowledge-based systems. Knowledge-based systems follow clearly defined instructions, such as IF-THEN rules that evaluate data to determine the appropriate response or action. Non-knowledge-based systems are not programmed to follow set rules, instead utilising AI, ML or statistical pattern recognition [3]. AI refers to the ability of machines or computers to simulate intelligent behaviour. It also refers to software that performs tasks or produces output that was previously thought to require human intelligence [4]. ML is therefore a subset of AI that is not explicitly programmed to follow rules, with computers having the ability to learn and adapt by analysing patterns in data using algorithms and statistical models [5].

ML has been explored across a range of clinical functions in emergency care for children, with some studies focusing on its use in triage and risk stratification, such as predicting clinical outcomes at triage [6] and predicting interventions for suspected sepsis [7]. For diagnostic support, ML has been applied to medical conditions such as appendicitis [8] and fractures [9]. Other research has examined ML for treatment-related decision support (such as anti-microbial tools that assist clinicians to safely switch patients from intravenous to oral antibiotics [10]) and operational efficiency (such as forecasting overcrowding [11]). However, when Chan et al. [12] carried out a scoping review on implementation of prediction models in EDs, only 3/31 of the implementations identified used ML. The applications used included sepsis, admission (inpatient and ICU) and deterioration due to acute coronary syndrome prediction. Most tools used in practice were scoring systems derived from existing clinical risk scores, clinical expertise, or logistic regression coefficients. This disconnect between AI/ML innovation and implementation mean it is essential to understand frontline perspectives on ML for decision support, before future effective and successful implementation can be achieved.

The objective of this study was to assess the understanding of key AI concepts, barriers in adopting ML based tools, current experience and opinions on the potential application of these tools for children’s emergency care staff (doctors, nurses and support staff) in the United Kingdom and Ireland. By capturing perspectives across various roles and experience levels, this study aimed to identify knowledge gaps, training needs, and factors that may influence successful integration of ML into children’s emergency care workflows.

## Materials and methods

An online voluntary anonymised survey study was carried out, with questions (S1 File) split into seven sections informed by previous research [13–18] and refined through discussion amongst the study investigators (Table 1). The survey was designed in Research Electronic Data Capture (REDCap) [19,20], a secure online software platform. Branching logic was used to minimise the total number of questions presented to the respondents. For sections containing numerous questions, random ordering was used to minimise respondent fatigue and improve the quality of responses. All (apart from two qualitative questions) were mandatory. The anonymised dataset, accessible only to the study investigators, was stored on a secure server in Technological University (TU) Dublin.

**Table 1.** Survey sections, with number of questions and answer types.

| Survey Section | Number of questions | Types |
| --- | --- | --- |
| Study participant information | 6 | Multiple choice, single answer |
| Participant demographics | 5-10 | Multiple choice, drop-down list<br>Text<br>Multiple choice, Single answer<br>Multiple choice, multiple answer |
| Confidence in understanding key artificial intelligence concepts | 7-13 | Likert (5 values)<br>Text<br>Multiple choice, single answer |
| Current experience with clinical decision support system tools | 4-8 | Multiple choice, Single answer<br>Text<br>Multiple choice, multiple answer |
| Reasons machine learning decision support tools are not used in their clinical setting | 0-10 | Likert (5 values)<br>Text |
| Training in machine learning | 3-4 | Multiple choice, Single answer<br>Text<br>Multiple choice, multiple answer |
| Areas of application for machine learning in emergency medicine | 24-25 | Likert (5 values)<br>Text |

Participants were presented with six questions assessing their level of understanding of key AI concepts. They were then asked to watch a short video (3.5 minutes) embedded within the survey explaining key AI concepts and providing examples of decision support tools (https://vimeo.com/997910349). After the video they were asked if they would like to change their original answers, and if they answered yes, the six questions were repeated. The survey was tested for usability and technical functionality by the study investigators and an independent clinician, with revisions made based on feedback.

The survey was distributed through Paediatric Emergency Research in the United Kingdom and Ireland (PERUKI) [21]. Sites comprised a variety of types and locations, including standalone paediatric and mixed adult-paediatric emergency EDs in both urban and rural settings. An email was sent to each of the 75 PERUKI site leads containing a link to the survey and instructions. Given variability in site size (and consequent staffing levels), site leads were provided with guidance on minimum and maximum numbers of invites by staff group, and communicated to the study team how many individuals had been invited in order to calculate response rates:

- Doctor: minimum 5, maximum 10
- Nurse: minimum 5, maximum 10
- Support Staff: minimum 2, maximum 4

The survey was open from 29th August 2024 until 13^th^ October 2024. Three reminders were sent to site leads; the first was two weeks after opening, with the next two reminders two weeks and 72 hours before the survey ended.

Data were exported from REDCap and prepared for analysis by reformatting variables, removing all incomplete responses, generating categorical variables, and verifying data integrity using REDCap and Microsoft Excel. Most of the data analysis was performed in Microsoft Excel and is presented using descriptive statistics including counts, percentages, and visual representation in charts. Likert scale responses of ‘Agree’ and ‘Strongly Agree’ were grouped and reported together, as was ‘Likely’ and ‘Extremely Likely’ where applicable.

To assess whether years of experience in emergency medicine was associated with self-reported understanding of key AI concepts, chi-square tests to determine association were performed. Survey responses were grouped by participant type, AI concept, and years of experience. Each concept was coded with a binary outcome (‘Agree’ and ‘Other’). Significance was set at p <0.05, with pairwise comparisons performed using Fisher’s exact test with Bonferroni correction (significance at p < 0.0083) for post-hoc analysis. Odds ratios and adjusted p-values were reported. Analyses were performed in R (version 4.3.3). This study used the Checklist for Reporting Results of Internet E-Surveys (CHERRIES) guidelines [22] and the completed checklist is provided in S2 File.

### Ethical approval and participant consent

Ethical approval for this cross-sectional survey study was received from the Research Ethic Committees of both Children’s Health Ireland (REC-281-23) and TU Dublin (LEWS-023-83). Participants provided electronic written informed consent by affirming six mandatory statements regarding their understanding of the study and agreement to participate prior to gaining access to the survey (S1 File).

## Results

The overall survey response rate was 72.3% (660/913), 77.7% (328/422) for doctors, 71.2% (262/368) for nurses and 56.9% (70/123) for ED support staff. Most respondents were doctors at 49.7% (328/660), from England (75.5%, 498/660) and had six or more years’ experience in emergency medicine (60.0%, 396/660) (Table 2).

**Table 2.** Respondents Characteristics.

| Characteristic | Doctor<br>(n=328)<br>n (%) | Nurse<br>(n=262)<br>n (%) | Support Staff<br>(n=70)<br>n (%) | All<br>(n=660)<br>n (%) |
| --- | --- | --- | --- | --- |
| **Country/Constituent Country** |  |  |  |  |
| England | 251 (76.5) | 197 (75.2) | 50 (71.4) | 498 (75.5) |
| Republic of Ireland | 38 (11.6) | 34 (13.0) | 14 (20.0) | 86 (13.0) |
| Scotland | 23 (7.0) | 17 (6.5) | 1 (1.4) | 41 (6.2) |
| Wales | 9 (2.7) | 7 (2.7) | 2 (2.9) | 18 (2.7) |
| Northern Ireland | 7 (2.1) | 7 (2.7) | 3 (4.3) | 17 (2.6) |
| **Years working in Emergency Medicine** |  |  |  |  |
| 0-2 | 57 (17.4) | 49 (18.7) | 19 (27.1) | 125 (18.9) |
| 3-5 | 74 (22.6) | 51 (19.5) | 14 (20.0) | 139 (21.1) |
| 6-10 | 86 (26.2) | 68 (26.0) | 13 (18.6) | 167 (25.3) |
| 10+ | 111 (33.8) | 94 (35.9) | 24 (34.3) | 229 (34.7) |
| **Respondent Type (Top 6 for each Role)** |  |  |  |  |
| Consultant | 154 (23.3) |  |  |  |
| Specialty Registrar | 86 (13.0) |  |  |  |
| Senior House Officer | 27 (4.1) |  |  |  |
| Senior Fellow | 15 (2.3) |  |  |  |
| Junior Fellow | 15 (2.3) |  |  |  |
| Specialty and Associate Specialist | 12 (1.8) |  |  |  |
| Band 6 |  | 71 (10.8) |  |  |
| Band 5 |  | 62 (9.4) |  |  |
| Advanced Clinical Practitioner |  | 42 (6.4) |  |  |
| Band 7 (Sister/Charge Nurse) |  | 41 (6.2) |  |  |
| Clinical Nurse Manager 2 |  | 20 (3.0) |  |  |
| Advanced Nurse Practitioner |  | 8 (1.2) |  |  |
| Operational/Administrative Support |  |  | 33 (5.0) |  |
| Patient Flow/Bed Manager |  |  | 10 (1.5) |  |
| Pharmacist |  |  | 8 (1.2) |  |
| Healthcare Assistant |  |  | 4 (0.6) |  |
| Social Worker |  |  | 3 (0.5) |  |
| Play Specialist |  |  | 2 (0.3) |  |
| **Clinician Type** |  |  |  |  |
| Emergency Medicine | 123 (37.5) |  |  |  |
| Paediatrics with PEM subspecialty training | 95 (29.0) |  |  |  |
| Paediatrics | 57 (17.4) |  |  |  |
| Emergency Medicine with PEM subspecialty training | 46 (14.0) |  |  |  |
| Other | 7 (2.1) |  |  |  |

Prior to watching the video, the concept of ‘Artificial Intelligence’ was the most familiar at 60.3% (398/660), compared to the least known ‘Deep Learning’ at 19.1% (126/660). ED support staff had a greater initial understanding of all concepts, doctors were next apart from the concept ‘Natural Language Processing’ (doctors: 23.2% (76/328), nurses: 23.7% (62/262)). After watching the video 40.2% (265/660) changed their answers, with doctors changing most (43.3%, 142/328). The largest increase in understanding was for ‘Generative Artificial Intelligence’, increasing from 21.4% (141/660) to 44.8% (296/660), with doctors’ understanding increasing the most by 25.7% (Fig 1).

**Fig 1.**
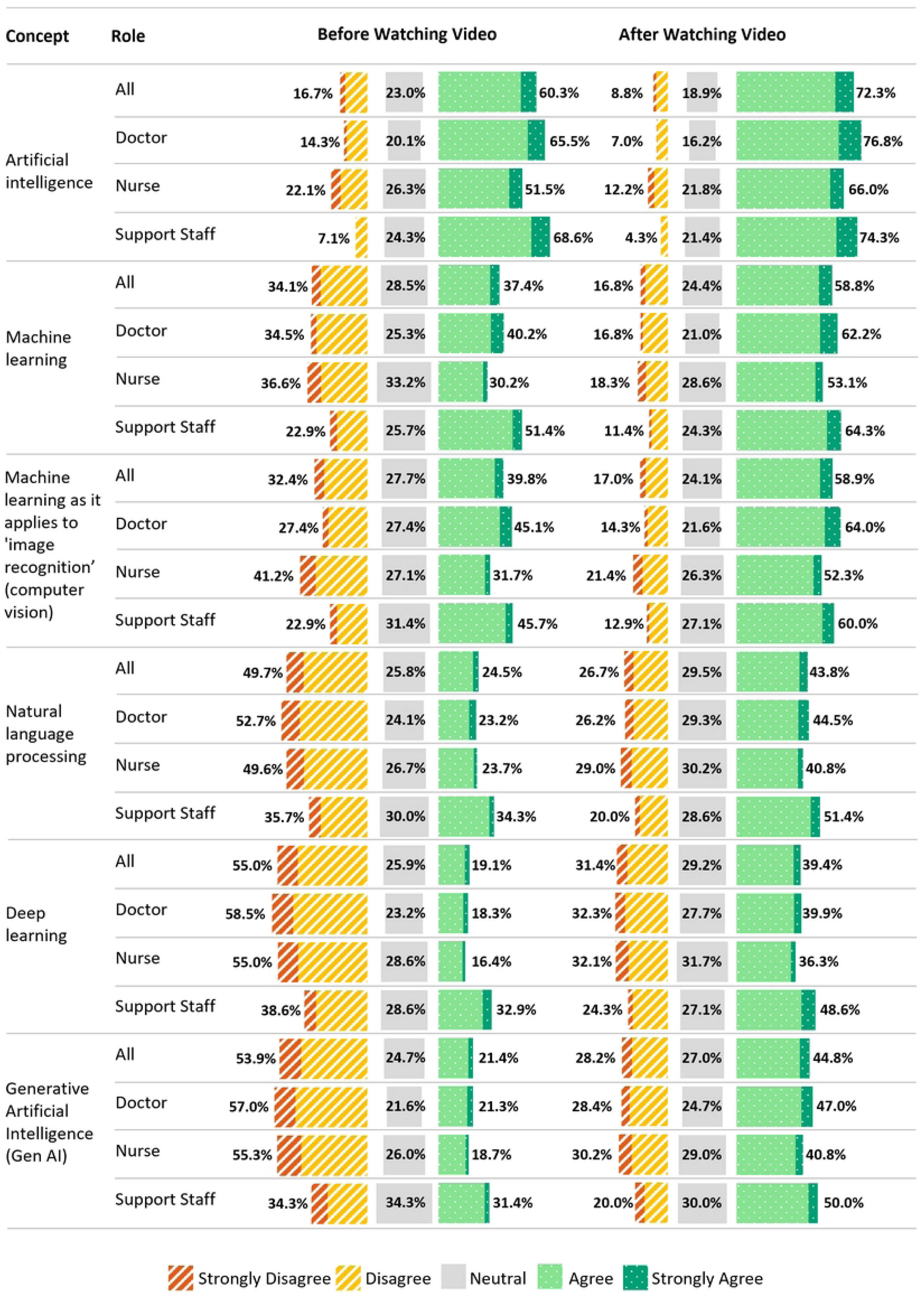
Confidence in understanding key artificial intelligence concepts by participant type, pre and post watching educational video.

The results of tests performed on the association between years of experience and self-reported initial understanding of key AI concepts, grouped by respondent type, revealed a significant association among nurses in their understanding of generative AI only (χ²(3) = 9.65, p = 0.0217). Nurses with 0–2 years of experience were more likely to report understanding generative AI compared to those with 3–5 years of experience (odds ratio = 4.39, 95% CI [1.36-16.91], adjusted p = 0.0396). No other comparisons between groups were statistically significant (S1 and S2 Tables).

Most respondents (73.6%, 486/660) advised that ML decision support tools were not used in their clinical setting (doctors: 79.9% (262/328), nurses: 67.2% (176/262), support staff: 68.6% (48/70)). For reasons why, some respondents had concerns around deciding which process would benefit the most (42.8%, 208/486), not having skilled resources to develop these tools (37.0%, 180/486), the explanatory capabilities of these tools not being well developed (31.5%, 153/486), and not enough or poor data quality (28.8%, 140/486). On the positive side, many respondents agreed that these tools can be integrated well into clinical workflows (60.7%, 295/486), would have trust in these tools (58.2%, 283/486), are convinced of the value for decision support (53.7%, 261/486), and where relevant, have not previously found them difficult to use (30.5%, 148/486) (Fig 2).

**Fig 2.**
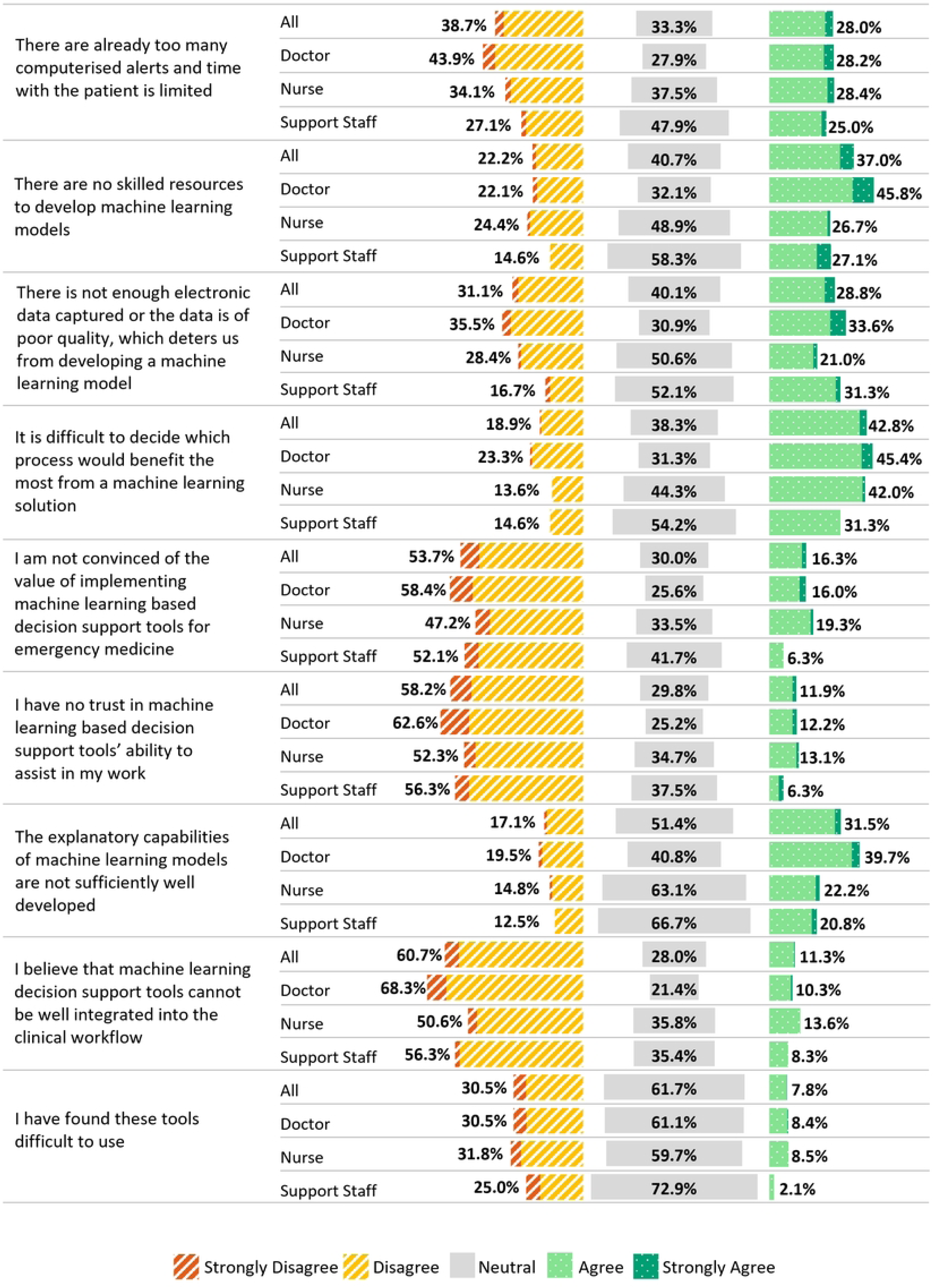
Opinion on why machine learning decision support tools are not used in clinical settings by participant type.

Use of knowledge-based tools was 53.3% (352/660) across all respondents, and much lower for non-knowledge-based (ML) tools at 7.7% (51/660), with more nurses than doctors using these tools (nurses: 9.9% (26/262), doctors: 6.4% (21/328)) (Table 3).

**Table 3.** Use of knowledge and non-knowledge-based tools.

| Participant Type / Answer | Knowledge based (rule based)<br>n (%) | Non-knowledge based (machine learning)<br>n (%) |
| --- | --- | --- |
| **All (n=660)** |  |  |
| Yes | 352 (53.3) | 51 (7.7) |
| No | 193 (29.2) | 609 (92.3) |
| Unsure | 115 (17.4) |  |
| **Doctor (n=328)** |  |  |
| Yes | 172 (52.4) | 21 (6.4) |
| No | 105 (32.0) | 307 (93.6) |
| Unsure | 51 (15.5) |  |
| **Nurse (n=262)** |  |  |
| Yes | 155 (59.2) | 26 (9.9) |
| No | 53 (20.2) | 236 (90.1) |
| Unsure | 54 (20.6) |  |
| **Support staff (n=70)** |  |  |
| Yes | 25 (35.7) | 4 (5.7) |
| No | 35 (50.0) | 66 (94.3) |
| Unsure | 10 (14.3) |  |

In relation to applications that would benefit the most from a ML approach to decision support, early warning of clinical deterioration ranked highest (83.4%, 492/590), with both doctors (84.5%, 277/328) and nurses (82.1%, 215/262) agreeing this was the most impactful use case. Ranked joint highest for doctors was analysis of radiology images (84.5%, 277/328). The lowest ranking application for doctors and nurses was diagnosis of mental health conditions (doctors: 19.2%, 63/328, nurses: 25.2% (66/262)) (Fig 3). When asked what additional applications could benefit from a ML approach, a small number (5.6%, 37/660) suggested additional applications such as workforce and rostering, patient flow, coding and discharge summary/correspondence, and medication safety/management. These applications were jointly ranked highest at 5/660 (S1 Fig).

**Fig 3.**
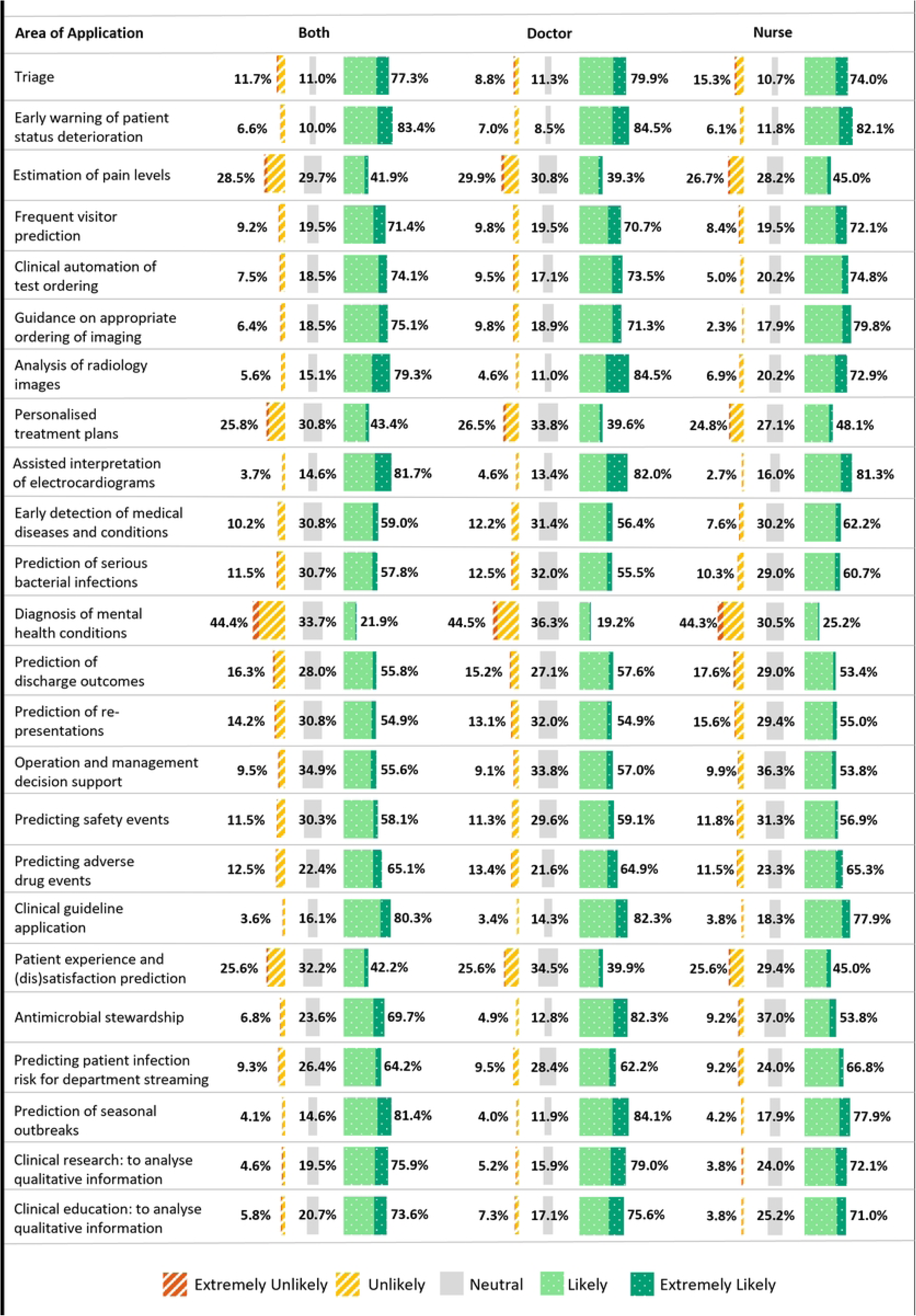
Areas of application of machine learning for decision support by participant type (doctor and nurse).

Clinically focused ED support staff (nurses, paramedics, pharmacists, scientists and health and social care professionals) ranked clinical automation of test ordering and prediction of seasonal outbreaks highest (71.0%, 22/31) and provided the largest proportion of neutral responses for predicting patient infection risk for department streaming (54.8%, 17/31). Non-clinical ED support staff (administrators, healthcare assistants) ranked clinical guideline application highest (66.7%, 26/39) and were most neutral about anti-microbial stewardship (69.2%, 27/39). Both groups ranked diagnosis of mental health conditions lowest (clinical: 25.8%, 8/31; non-clinical: 23.1%, 9/39) (Fig 4).

**Fig 4.**
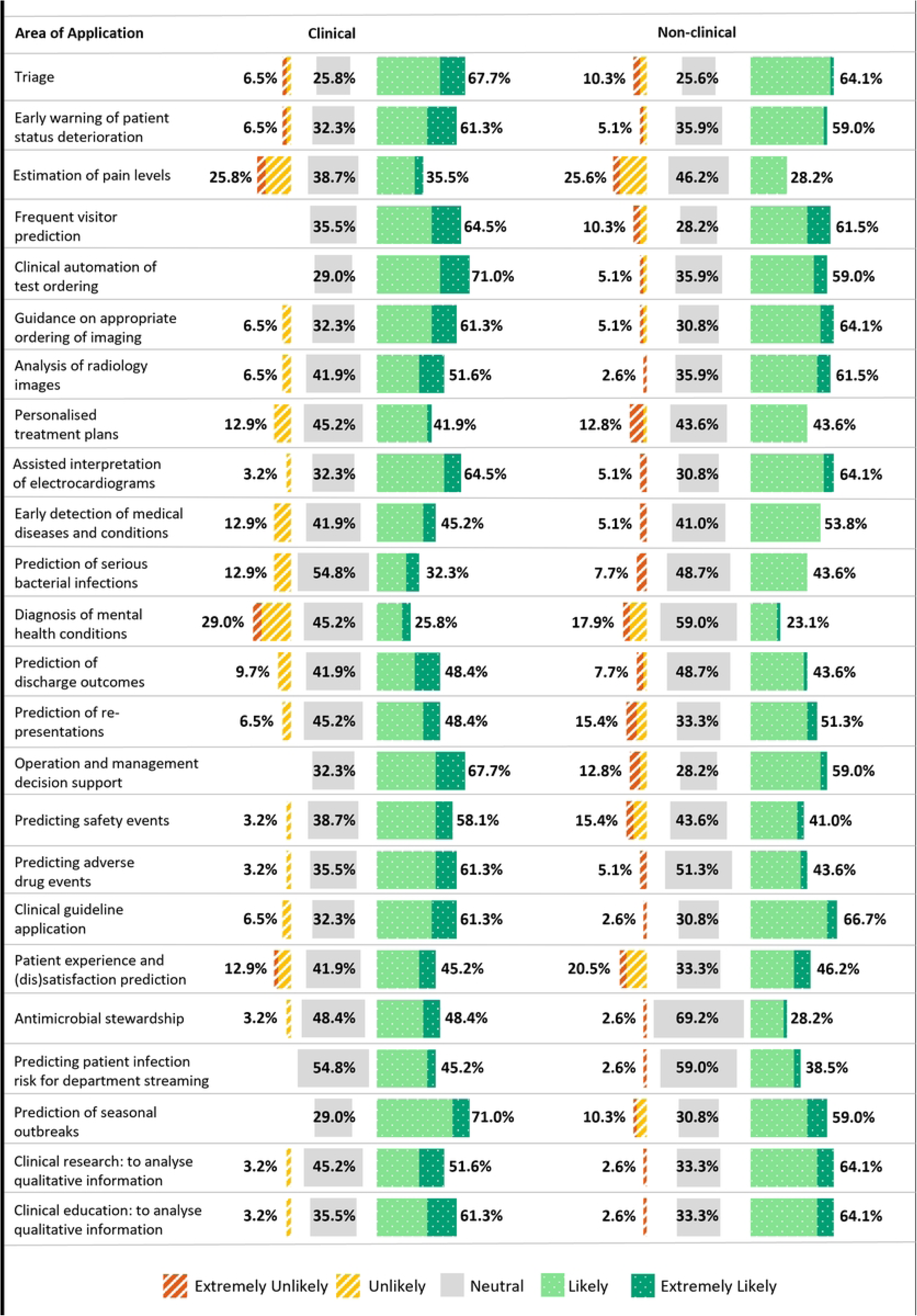
Areas of application of machine learning for decision support by role type grouping (support staff)

When respondents were asked if they had contributed to a project that used ML in emergency medicine, 9.1% (60/660) had, most of whom were nurses (32/660). Few had received training in ML (7.6%, 50/660) and even less had accredited training (2.9%, 19/660). However, there was a strong interest in furthering their knowledge in ML (78.2%, 516/660) (Table 4).

**Table 4.** Training and experience with machine learning.

| Question and Response | All<br>(n=660)<br>n (%) | Doctor<br>(n=328)<br>n (%) | Nurse<br>(n=262)<br>n (%) | Support Staff<br>(n=70)<br>n (%) |
| --- | --- | --- | --- | --- |
| **Have you contributed to a project that used machine learning in emergency medicine?** |  |  |  |  |
| Yes | 60 (9.1) | 20 (6.1) | 32 (12.2) | 8 (11.4) |
| No | 600 (90.9) | 308 (93.9) | 230 (87.8) | 62 (88.6) |
| **Have you received any training in machine learning?** |  |  |  |  |
| Yes | 50 (7.6) | 14 (4.3) | 31 (11.8) | 5 (7.1) |
| No | 610 (92.4) | 314 (95.7) | 231 (88.2) | 65 (92.9) |
| **What type of training?** |  |  |  |  |
| Accredited | 19 (2.9) | 6 (1.8) | 11 (4.2) | 2 (2.9) |
| Non-accredited | 13 (2.0) | 2 (0.6) | 9 (3.4) | 2 (2.9) |
| Casual | 18 (2.7) | 6 (1.8) | 11 (4.2) | 1 (1.4) |
| **Do you have an interest in furthering your knowledge in machine learning?** |  |  |  |  |
| Yes | 516 (78.2) | 259 (79.0) | 201 (76.7) | 56 (80.0) |
| No | 144 (21.8) | 69 (21.0) | 61 (23.3) | 14 (20.0) |

## Discussion

This study assessed the understanding of key AI concepts, the barriers in adopting ML based tools, current experience and opinions on the potential application of these tools for paediatric emergency care amongst the wider ED workforce in the United Kingdom and Ireland. The volume of responses and response rate was strong at 660 and 72% respectively. There was representation across participant types, roles and experience. Initial familiarity with AI concepts varied with AI being the most well-known concept compared to more specialised areas such as deep learning. Years of experience working in or with emergency medicine was generally not a significant factor in understanding these AI concepts across the participant types, except for a single finding among nurses related to generative AI. However, this isolated result among nurses may be attributable to chance alone, given the number of questions asked. Respondent understanding of concepts improved after watching a short video, particularly regarding generative AI, with the largest change seen among medical staff. ML decision support tools were not widely used, with reasons including not knowing which process would benefit the most, lack of skilled resources, inadequate explanatory capabilities, and poor data quality. Most respondents agreed these tools could be integrated into clinical workflows and expressed trust in their use for decision support. Respondents ranked early warning systems highest for promising ML based applications; diagnosis of mental health conditions was lowest. Actual involvement in ML projects and training was rare, however interest in furthering knowledge was high. These results demonstrate both uncertainty and enthusiasm within the ED workforce for paediatric ML based tools, emphasising the need for more targeted education, clearer implementation plans, improved explainability and trustworthiness to support adoption and greater exposure to ML tools for use within ED clinical workflows.

### Knowledge and training

The wider ED workforce had a lower baseline understanding of AI concepts before watching the video compared to a previous similar survey completed by digital/PERUKI site leads, consisting of 65 responses in the same setting (96.9% of whom were consultants) [23] which lends face validity to the observed differences. In the site leadership survey 80.0% and 32.2% understood the concepts of ‘Artificial Intelligence’ and ‘deep learning’ respectively, compared to 60.3% and 19.1% from the wider ED workforce. Site leadership may have a better understanding due to their leadership position and may be more involved in discussions in research or strategic planning for AI or ML decision support tools. Clear from both survey results was the particularly low understanding of deep learning, enforcing the need for AI literacy efforts to cover not just basic concepts, but also key techniques used in modern clinical tools, such as deep learning models for sepsis that are already improving outcomes in emergency care settings [24].

In a survey of healthcare professionals (medical doctors, nurses, therapists and others), Castagno et al. [25] demonstrated that half the respondents were not familiar with the concepts of ML and deep learning. Since that UK survey was performed in 2020, adoption of AI into medicine appears to be slow despite rapid advances in AI, as we have shown a similar lack of understanding of AI concepts amongst healthcare professionals in 2024. This may be partly due to a lack of technological understanding among staff [25], and education is therefore key to better adoption of these tools in clinical practice. The European Union (EU) AI act has reinforced this need by requiring providers and deployers of AI systems ensure their staff have sufficient AI literacy to use these tools. The act defined AI literacy as understanding, skills and knowledge needed to make informed decisions on the effective implementation of AI systems, along with gaining awareness of the benefits, risks and potential to cause harm [26]. Kimiafer et al. [27] carried out a systematic review on AI literacy among students and healthcare providers, revealing findings similar to ours in that few healthcare professionals demonstrated adequate AI literacy or had received training. The researchers found that amongst healthcare professionals, radiologists had the highest level of AI literacy compared to the other specialists. Nevertheless, there was a shared motivation across all groups to adopt AI to enhance healthcare delivery.

Although few had received training, there was also a strong interest from the respondents in our survey in furthering their AI literacy. To help medical professionals confidently adopt and recommend AI tools, building AI literacy through comprehensive education and training is essential [28]. It is however essential to recognise that training may need to be customised by role; for example, site leaders may require more advanced AI training, enabling them to better lead AI adoption and make strategic implementation decisions compared to the wider ED workforce whose training may be more practical in nature. This may focus on the basics of understanding AI and ML, interpreting outputs responsibly, and learning how to integrate these tools into workflows. Misra et al. [29] support this view of tailored AI training, grouping training needs into three types of learning; all clinicians (foundational understanding), most clinicians (product specific training), and some clinicians acting as clinical champions (advanced domain specific training in order to create and promote ideas for the safe, equitable and effective use of AI within their clinical domain).

### Barriers to ML adoption

One of the key challenges of implementing ML for decision support reported is determining which workflow or problem would benefit the most. This viewpoint is confirmed in the literature, through suggestion that AI implementation should start with a clear problem as opposed to the analysis of data in search of a solution [30]. Without a clearly defined problem, staff may remain hesitant in championing and adopting these tools. Lack of explainability in how ML decision support tools produce output was also a concern. Researchers have previously found that the main barrier to adoption was model explainability and interpretability, with deep learning models often viewed as black boxes, unlike more transparent and traditional methods like logistic regression [31]. Policy initiatives including the EU AI act mandate strict requirements for high-risk AI systems, calling out transparency and human oversight among other factors to ensure the protection of fundamental rights and potential safety issues posed by AI adoption [26]. Our survey results and the literature emphasise how crucial explainability is for adoption, with trust in these tools depending on clear evidence, and either interpretability or robust validation to gain acceptance for use of these tools in emergency care.

Another concern was the lack of skilled resources to develop ML models. Co-design and collaborative development is essential for the successful implementation of these tools, typically involving a range of stakeholders/skilled resources. These include a principal investigator or project lead who understands the healthcare setting and organisational context, data scientists with expertise in model development and design, ML engineers responsible for deploying the model, clinicians or users who interpret and apply the outputs, data engineers or information technology experts who understand the source data and complexities and inconsistencies of electronic health record systems [32]. Sendak et al. [33] integrated a sepsis deep learning model into ED clinical care which was delivered by a multidisciplinary team consisting of clinicians, data scientists, data engineers and statisticians. This collaborative approach not only facilitated the development of the tool but ensured clinical relevance, usability, and trust amongst end-users. The World Health Organization (WHO) stated that AI technologies should not be designed solely by scientists and engineers, but through inclusive, transparent processes that engage end-users and stakeholders from the outset [34]. However, such technical expertise needed for these collaborations may not be available in all organisations, highlighting the value of partnerships with universities, research institutions or industry. In our previous study of digital/PERUKI site leadership, 69.2% were concerned about the lack of skilled resources [23] compared to 37.0% in this survey. This increased concern from the digital/PERUKI site leadership may reflect a clearer understanding of the technical and workforce challenges involved in deploying these tools in practice. Site leaders were also more concerned about data quality issues (55.8%) compared to the wider ED workforce (28.8%).

The performance of any ML model is constrained by the quality of the underlying data [35]. The WHO [36] warns that poor quality or missing data can result in AI models learning misleading patterns in the data, whereby artefacts may be predicted instead of actual clinical outcomes. This could limit the effectiveness of these tools, and due to insufficient usable data, these models may not be available for specific populations. The European Union AI act [26] goes a little further stating that if an AI system is not trained on high-quality data or properly tested for accuracy and robustness, it may single out people in a discriminatory, incorrect, or unjust manner. Researchers acknowledge that in a paediatric emergency medicine context there is a paucity of comprehensive datasets, a known barrier to the widespread adoption of AI [37]. Recognition of the importance of data quality to ML has seen the adoption of standards and quality frameworks [38]. Data quality dimensions have emerged to address the issue, including accuracy, completeness, consistency, reasonability, validity and uniqueness/de-duplication [39]. Encouragingly, the wider ED workforce and particularly the leadership from our previous survey [23] recognised the importance of high-quality data, confirming an awareness of the challenge this brings in developing and maintaining quality ML tools.

To address the concerns and barriers to adoption of these ML based tools, there is a clear and urgent need for AI governance frameworks. Governance frameworks should have trust at its centre, integrating principles such as fairness, ethics, accountability, transparency and safety, along with aligning with societal and organisational standards to guide the full lifecycle of AI system integration in practice [40]. Hassan et al. [40] also emphasise the need for stakeholder engagement within the full lifecycle (initiation to deployment and ongoing monitoring).

### Exposure to ML tools in a clinical setting

There was minimal exposure to ML decision support tools in clinical settings. This is perhaps unsurprising, as despite voluminous research into the utility of AI and ML decision support tools in healthcare, few have been deployed in real-world settings [41]. Many respondents had used knowledge (rule) based systems, but given the limited understanding of AI technology, some may not have recognised that these knowledge-based tools were powered by ML.

This limited exposure and uncertainty around ML tools among respondents reflects broader challenges identified in the literature. Peek et al. [42] conducted a review of the literature on the implementation of AI-based clinical decision support systems (CDSS), finding it to be an emerging and underdeveloped area of research. Several of their findings align with ours. These include the lack of well-developed explanatory capabilities, which relates closely to concerns around transparency and trust in their review, and the limited AI knowledge among healthcare professionals. Post-implementation, the researchers identified poor adoption linked to dissatisfaction with accuracy, poor system integration, and limited perceived usefulness of the tools outputs. Similarly, our survey highlights the difficulty in determining which workflows would benefit the most from ML-based decision support. The authors made several recommendations, including the importance of grounding AI-CDSS implementation research in established frameworks from implementation science to ensure consistency and comparability across studies. They also emphasised the need to explore the perspectives of broader stakeholder group such as patients, carers, the public, leaders and healthcare managers, while suggesting that healthcare professional’s perspectives should be examined post-implementation. Finally, they called for more real-world implementation studies to capture human, technical, and organisational challenges, and recommended investment in reusable infrastructure for the validation and deployment of these tools, mitigating the need for major infrastructural changes.

### Enablers to ML adoption

The digital/PERUKI site leadership from our previous study had a higher confidence in trusting ML decision support tools [23] compared to the wider ED workforce (Leads: 76.9%; Wider ED workforce: 58.2%). Trust in these tools is linked to perceived understandability and accuracy, essential for ML decision support adoption [43]. The results of our previous survey showed that leadership had a greater understanding of the AI key concepts they were presented with, indicating the likelihood of greater exposure or research into these tools. We are again brought back to the importance of transparency, all models intended to interact with natural persons in healthcare must provide transparency information for downstream providers [44]. To foster trust in these tools, there are known requirements that must be realised to enable trustworthy AI, these include transparency, human agency and oversight, privacy and data governance, accountability, technical robustness and safety, environmental and societal well-being and diversity, non-discrimination and fairness [45].

Most respondents believed these tools can integrate well into clinical workflows. Researchers have found that successful AI implementation is the ability to integrate into clinical workflows, ideally embedding them into existing hospital information systems and streamlining their use to avoid adding additional steps. It is therefore important that end-users are engaged early in the process from development, right up to implementation, guiding the most efficient way of integrating into existing workflows [31]. Implementation of these tools in practice, such as a study in ML Sepsis early warning system, tells us that even well-performing models can face resistance if they disrupt established clinical workflows or lack explainability. Incorporating feedback loops and minimising workflow interruptions were recognised in their study as factors to ensure successful adoption [46].

### Potential applications

Early warning of clinical deterioration was ranked highest by doctors and nurses for potential ML based decision support tools. Research into ML based early warning systems has shown great potential [47]. In one paediatric study, a deep learning early warning system was developed which outperformed the modified paediatric early warning score in accurate early prediction of deterioration events in any clinical situation, with fewer false alarms and a reduced number of cases to review [48]. Triage can function as an early warning system, with a recent prioritisation exercise carried out by the James Lind Alliance [49] including triage in the top 10 priorities for paediatric emergency medicine research. Triage was also one of the few areas in this survey where nurses demonstrated more certainty, with only 10.7% selecting a neutral response, compared to their greater hesitancy across all other applications. There is potential for ML to assist with patient triage, one type of approach to patient prioritisation was to predict outcomes such as hospitalisation, critical care and mortality [50]. Another approach is to predict the triage category or emergency severity index itself [51]. A narrative review carried out by Da’Costa et al. [52] on AI driven triage tools outlined that along with enhancing the accuracy of patient prioritisation, there is the potential to reduce wait times and improve resource allocation in the ED. These tools also provide consistency in triage decision-making and may help to reduce clinician cognitive burden. Although many of these ML tools are still in the research phase, there is potential for these ML driven alerts to assist with prioritising at risk patients, improving responsiveness and reducing alarm fatigue.

Doctors also selected the analysis of radiology images as a top use case, potentially due to a recognition of how AI is improving efficiency, accuracy and personalised healthcare in diagnostic imaging [53]. Some of the AI tools currently in use for ED radiology imaging include fracture detection, the classification of intracranial haemorrhage into poor and good prognosis, anomaly detection in chest radiographs, and opportunistic screening [54]. However, it has been recognised that there is a disparity in AI development and market availability between paediatrics and adults, with challenges reported on attempting to generalise adult algorithms to a paediatric population. These challenges include age related differences in anatomy, physiology, diagnoses, and imaging techniques [55]. Sammer et al. [55] also outlines what should be done to progress the development of safe and effective AI tools for children, including developing models with paediatric data, promoting collaboration across institutions and industries, enhancing transparency through regulatory frameworks, prioritising funding for paediatric AI research, and ensuring equitable access and appropriate clinical oversight specific to paediatric needs.

In our survey of PERUKI site leaders, anti-microbial stewardship was ranked highest at 90.8% [23]. In this current survey, doctors still ranked it highly (82.3%) but only 53.8% of nurses considered it likely to benefit from ML, suggesting leaders have greater understanding of the strategic importance of anti-microbial stewardship. For admitted patients, neither group tends to engage a lot with stewardship beyond completing the required documentation, the admitting team would have greater involvement. Nurses may also have ranked ML applications higher for areas that are more central to their role such as early warning systems. Researchers have recognised the need to advance anti-microbial stewardship techniques in paediatric emergency medicine utilising AI and ML methods [56], however although there has been lots of research into this area, a key challenge identified by researchers is the lack of evidence supporting the long-time effectiveness of AI for anti-microbial stewardship in real world scenarios [57].

Overall, ED support staff most frequently selected neutral responses when asked about potential ML applications, which is not unexpected given that many of these potential tools fall outside the scope of their roles. For example, when asked about antimicrobial stewardship, 48.4% of clinical and 69.2% of non-clinical support staff responded neutrally. Clinical support staff (which includes bed managers) showed strong support for ML applications that enhance operational tasks such as clinical automation of test ordering and prediction of seasonal outbreaks (both at 71%), which suggests a recognition of how ML could streamline workflows and support resource or capacity planning. While non-clinical support staff ranked clinical guideline application highest at 66.7%, which may have been selected due to the perceived alignment with their role in adhering to policies, procedures, protocols and guidelines. Interestingly all participants including support staff ranked diagnosis of mental health conditions lowest.

### Comparison of views across the wider ED workforce

Nurses reported the highest usage of ML tools in their clinical setting and the greatest participation in ML-related projects, although much of their training was informal. They were also the least convinced about the value of implementing ML for decision support. Doctors were the most likely to change their answers on understanding AI concepts after viewing the educational video, they showed a stronger trust in ML, and a greater belief that these ML tools could be integrated into clinical workflows, though they were more concerned about limited explanatory capabilities, and a lack of skilled resources to develop ML models. Support staff demonstrated greater initial familiarity with AI concepts but expressed the most uncertainty around the explanatory capabilities, what tools could benefit from a ML approach and the availability of resources to develop these tools; however, this group had the greatest interest in furthering their knowledge in ML.

### Limitations

The accuracy of the results are dependent on self-reported responses in this survey that potentially could have led respondents to under or overestimate their understanding and experience with these tools. The use of a short educational video may have introduced variability in the interpretation of questions the respondents were presented with depending on their level of engagement and learning, however one of the aims of the video was to assist the participants in understanding the questions. Although the survey included responses of a broad workforce across the UK and Ireland, it may not be reflective of settings with different healthcare systems, infrastructure, resources and experience with AI.

## Conclusion

This study provided an overview of the varied understanding, limited exposure and cautious optimism for ML based decision support tools across the UK and Ireland for children’s emergency care. The most promising applications emerging from this survey were early warning systems and radiology applications, however concerns around model explainability, data quality and clinical relevance remained. Lack of training and fundamental knowledge of AI was apparent. To support the safe and effective implementation of ML tools, tailored role-based training needs to be provided, along with investment in infrastructure and electronic healthcare record systems that facilitate better data standardisation and data quality. Making the transition from research to real-world impact will require practical, transparent, and user-centred implementation strategies. Future research should focus more on the real-world application of these ML tools into paediatric emergency care clinical workflows.

## Data Availability

The data are not publicly available due to confidentiality and anonymisation commitments made to participants as outlined in the study's ethical approval. We are able to share aggregated results upon reasonable request.

## Supporting information

**S1 Table.** Association between years of experience and the self-reported understanding of key AI concepts, grouped by respondent type.

**S2 Table.** Post-hoc analysis of Nurses association between self-reported agreement by years of experience for the concept generative artificial intelligence.

**S1 Fig.** Additional applications proposed by respondents by participant type.

**S1 File. Survey Questionnaire.** Details of study participant information and the survey questions.

**S2 File. Survey Checklist.** Checklist for Reporting Results of Internet E-Surveys (CHERRIES).

## Acknowledgements

The authors would like to acknowledge Stewart McKenna, Children’s Health Ireland at Tallaght for carrying out survey usability and technical testing. We are also grateful to the research and innovation office in Children’s Health Ireland for providing access to develop the survey in REDCap and technical administrator support: Kenny Lynch, Tania Bautista and Juliusz Filipowski. Also acknowledged are the PERUKI site leads who distributed and for some sites also completed the survey: Meriel Tolhurst-Cleaver, Alder Hey Children’s Hospital NHS Foundation Trust, Liverpool; Daniel Murrell, Bedfordshire Hospitals NHS Foundation Trust - Luton and Dunstable University Hospital; Jonathan Adamson, Birmingham Children’s Hospital; Charlotte Munday, Bristol Royal Hospital for Children; Katherine Thompson, Chelsea and Westminster NHS Foundation Trust; Michael Barrett, Children’s Health Ireland at Crumlin; Sheena Durnin, Children’s Health Ireland at Tallaght; Patrick Fitzpatrick, Children’s Health Ireland at Temple Street; Emma Fauteux, Cork University Hospital; Hannah Walsh, Derriford Hospital, Plymouth; Darren Ranasinghe, Epsom General Hospital; Slyvester Gomes, Evelina London Children’s Hospital; Patrick Aldridge, Frimley Park Hospital; Mark Anderson, Great North Children’s Hospital, Newcastle Upon Tyne; Phil Peacock, Great Western Hospital, Swindon; Simon Richardson, Hull Royal Infirmary; David Hartin, Ipswich Hospital; Arshid Murad, James Cook University Hospital, Middlesbrough; Nicholas Richens, John Radcliffe Hospital, Oxford; Rachael Mitchell, King’s College Hospital London; Atif Latif, Kingston Hospital NHS Foundation Trust; Alice Downes, Leeds General Infirmary; Shane Fitzgerald, Leicester Royal Infirmary; Claire Kirby, Newham Hospital, London; Edward Snelson, Norfolk & Norwich University Hospitals; Neha Jain, North Middlesex Hospital; Pete Figg, Northern Devon Healthcare NHS Trust; Lee Tubman, Northumbria Healthcare NHS Foundation Trust; Paul Tanto, Northwick Park Hospital; Christopher Gough, Nottingham University Hospitals NHS Trust; Sharryn Gardner, Ormskirk & District General Hospital; Alan Charters, Queen Alexandra Hospital, Portsmouth; Gareth Patton, Royal Aberdeen Children’s Hospital; Michaela Lazner, Royal Alexandra Children’s Hospital, Brighton; Tom Waterfield, Royal Belfast Hospital for Sick Children; Manish Thakker, Royal Berkshire NHS Foundation Trust; Graham Johnson, Royal Derby Hospital; Hannah Stewart, Royal Devon and Exeter Hospital; Shye Wei Wong, Royal Free Hospital, London; Jen Browning, Royal Hospital for Children & Young People, Edinburgh; Steven Foster, Royal Hospital for Children, Glasgow; David Kung, Royal Manchester Children’s Hospital; Kirsty Challen, Royal Preston Hospital; Lorna Bagshaw, Royal Wolverhampton NHS Trust; Stephen Davies, Salisbury NHS Foundation Trust; Adrian Marsh, Shrewsbury & Telford NHS Trust; Esther Wilson, Somerset Foundation Trust Musgrove Hospital; Niall Mullen, South Tyneside & Sunderland NHS Foundation Trust; Alasdair Moffat, Southampton Children’s Hospital; Ellie Day, Southmead Hospital, North Bristol Trust; Heather Jarman, St George’s University Hospitals NHS Foundation Trust; Vanessa Merrick, St Mary’s Hospital, Imperial College Healthcare NHS Trust; Greg Cranston, The Grange Hospital, Newport; Emre Basatemur, The Royal London Hospital; Holly Brooker, Torbay and South Devon NHS Foundation Trust; Tulsi Patel, University College London Hospital; James Foley, University Hospital Galway; Marylyn Emeda, University Hospital Lewisham; George Simpson, University Hospital of North Tees; Michael Fox, University Hospital of Wales, Cardiff; Tadgh Moriarty, University Hospital Waterford; Katherine Priddis, Watford General Hospital; Rachel Shute, West Suffolk; Davin Amin, Wexham Park Hospital; Amutha Anpananthar, Whipps Cross Hospital, London; Alex Brown, Barnet Hospital; Catherine Williams, Bolton NHS Foundation Trust

